# Prevention and Control of Acute Respiratory Viral Infections in Adult Population: A Systematic Review and Meta-Analysis on Ginseng-Based Clinical Trials

**DOI:** 10.1101/2021.07.23.21260970

**Authors:** Frank Adusei-Mensah, Richard Osei Agjei, Luqman Oluwaseun Awoniyi, Lekpa K. David, Fatima Badmus Awoniyi, Oluwafikayo S. Adeyemi, Adedayo Olawuni, Ayobami Adegbite

## Abstract

**Introduction:** Acute respiratory infections are continuously emerging. Discovered in Wuhan city, China in 2019, COV-SARS-2 and most viral respiratory diseases presently do not have a definitive cure. This paper aims to evaluate the therapeutic effectiveness of ginseng for prevention and control of acute respiratory illness including SARS-COV-2 in adult population.

**Method:** We performed a systematic literature review using databases PubMed, Medline, Scopus, Google Scholar, Web of Science, and Cochrane library from 1st through the 27th of April 2020. All related articles that reported the use of Ginseng in COVID-19 patients were included in this analysis. Screening was done by 2-independent researchers. The meta-analysis was performed using comprehensive meta-analysis package.

**Result:** 596 articles were retrieved for the time frame. After screening, 5 articles with RCTs outcomes relevant to the review were selected. Ginseng was found to be effective in the reduction of risk by 38 % and 3-days shorter duration of acute respiratory illness (ARI) in all trials than placebo.

**Conclusion:** As the world continues to race to find a cure, it is important to consider the use of ginseng which has been proven over the years to be effective in the treatment of acute respiratory illnesses. Further studies should however be conducted to determine the right dosage to improve efficacy and prevent adverse events.

**Funding:** This research did not receive any specific grant from funding agencies in the public, commercial, or not-for-profit sectors.

**Highlight:**

⍰ COVID-19 is very infectious ravaging the globe
⍰ Millions have been infected and hundreds of thousands lost their lives to COVID-19
⍰ Due to absence of vaccines, urgent search for vaccines and drugs is still underway
⍰ Ginseng has been useful in similar respiratory viral infections in the past
⍰ Current paper throws more light on the need to consider ginseng for COVID-19 control

## 1.0 Introduction

Coronaviruses (CoVs) are a large family of viruses that cause illness ranging from mild illness like the common cold to more severe diseases such as Middle East Respiratory Syndrome (MERS-CoV) and Severe Acute Respiratory Syndrome (SARS-CoV). Corona viruses are usually zoonotic diseases transferred from animals to humans. SARS-CoV is believed to have been transmitted from civet cats to humans and MERS-CoV from dromedary camels to humans ^1^. Novel coronavirus disease (COVID-19) is a new strain of a corona virus first discovered in Wuhan city in China on 12^th^ December 2019 where it is believed the first animal to human transmission took place. The WHO officially named the corona virus on the 11th, February 2020 as COVID-19 and it was categorized as pandemic on 11^th^ March by WHO ^2^. The disease has been reported in over 190 countries, over 2 million deaths and over 100 million confirmed cases according to WHO’s report ^2^. The impact of COVID-19 pandemic has been greatly felt in all areas of development. Signs including respiratory symptoms, fever, cough, shortness of breath and breathing difficulties have been associated with the disease. In more severe COVID-19 cases, infection can cause pneumonia, severe acute respiratory syndrome, kidney failure and death ^1, 3^.

To the best of our knowledge, currently there is no approved therapy to treat COVID-19 patients ^4, 5^. As emerging infectious diseases are on the rise, the search for alternative vaccines and therapies has never been more anticipated than now. It is best to learn from the known to the unknown, lessons and management of previous similar viral diseases can be of great asset towards the fight against the current COVID-19 pandemic and other emerging acute respiratory diseases. In previous corona virus epidemics and acute respiratory diseases, herbal and alternative medicines have played important roles both in the prevention and in the treatment of the diseases. For instance, Chinese medicinal approaches were used to prevent and treat severe acute respiratory syndrome (SARS) from the 2003 SARS-coronavirus disease ^6^. Herbal and Traditional Chinese medicines (TCM) have played critical roles in previous viral diseases including SARS-CoV, influenza A H1N1, influenza A H7N9 and COV-SARS viral diseases ^7, 8^. Traditional Chinese medicines have also been used to prevent and treat severe acute respiratory syndrome (SARS) and H1N1 coronavirus diseases ^8, 9^. Ginseng has been traditionally used in Asia for thousands of years to treat a variety of ailments. Ginsenosides, poly and oligosaccharides found in ginseng have been shown in various studies to enhance immune response against viral diseases ^9^. In other studies, standardized ginseng extract was shown to have preventive and therapeutic effect on influenza virus patients with lower incidence of influenza and stronger immune responses against the disease ^10^. Ginseng has been used either alone or combined with other herbs for treating chronic respiratory diseases ^11^ and upper respiratory tract infections ^12, 13^. Ginseng has also played critical role in the treatment of respiratory viruses which are a major cause of influenza-like viral illness (ILI) including coronaviruses with symptoms characterized by sudden onset of high fever (> 38°C), headache and cough ^14, 15^ similar to COVID-19.

As the world seek for answers on how soon probable therapies and vaccines could be developed to control the spreading and to treat the infected persons of respiratory infections, the use of ginseng should be much investigated. Especially in resource limited countries to prevent infection with COVID-19 and to treat mild COVID-19 cases. However, to the best of our knowledge its effectiveness in treating symptoms associated with COVID-19 has not be explored. Despite the *in vivo, in vitro* and clinical trials done on ginseng to show their safety and efficacy in preventing and treating viral respiratory infections and chronic respiratory diseases, it has not been considered as potential therapeutic and preventive agent against COVID-19. We aimed in the present study to explore the efficacy of ginseng in preventing and treating acute respiratory viral diseases and the potential of ginseng to serve as preventive or treatment alternative for COVID-19. Also, the active compounds of ginseng could serve as agents for COVID-19 drug design and development.

## 2.0 Background

**Figure 1:**
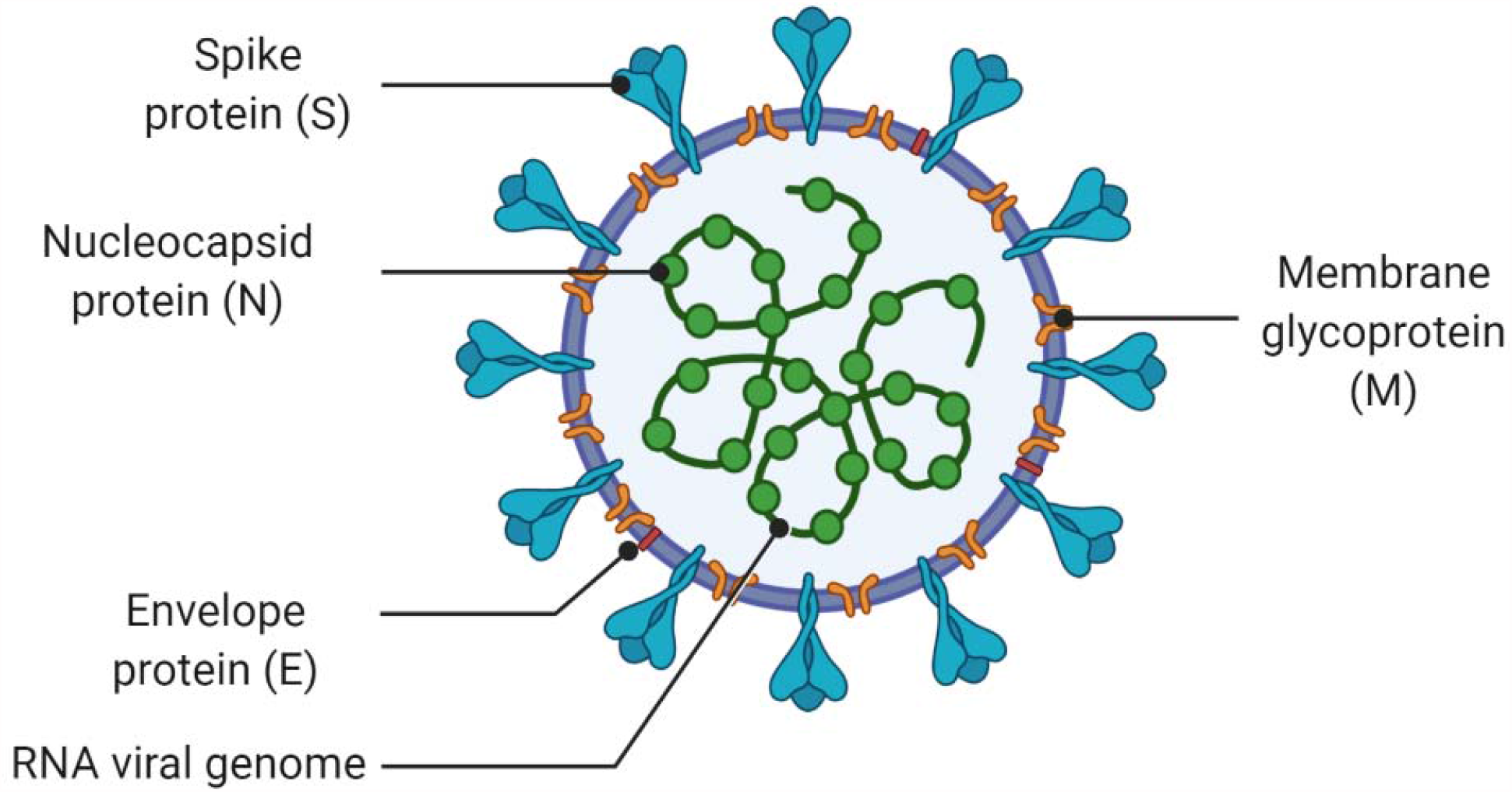
Pictorial view of COVID-19. (Created with BioRender)

### 2.1 Human Pandemic in Retrospect

It’s been eleven years since the world experienced its devastating last pandemic, the 2009 H1N1 swine flu. Within a space of a year specifically from spring 2009 through to spring 2010, the virus infected huge number of people totaling 1.4 billion across the world and claimed about 1 million lives ^16^. Presently, over 190 countries of the world have experienced the COVID-19 pandemic, caused by a novel coronavirus labelled SARS-CoV-2 with sequence homology to SARS-COV ^16(p19)^. The emergence of pandemic throughout history occurs at the human–animal interface, when animal infections (zoonotic infections) breach species barriers to infect human ^17^. During the past several centuries, pandemics that have been identified range from smallpox, measles, H1N1, influenza, AIDS, H2N2, H3N2, Spanish flu, and multi-drug resistance tuberculosis ^18, 19, 20, 21^). Table 1 gives more insight. As indicated earlier, the last pandemic the world saw is H1N1 swine flu which has several similar characteristics like the present COVID-19 in terms of rates of infection and mortality. Both past and present pandemics have cost the world trillions and trillions of U.S dollars ^22^.

**Table 1:**
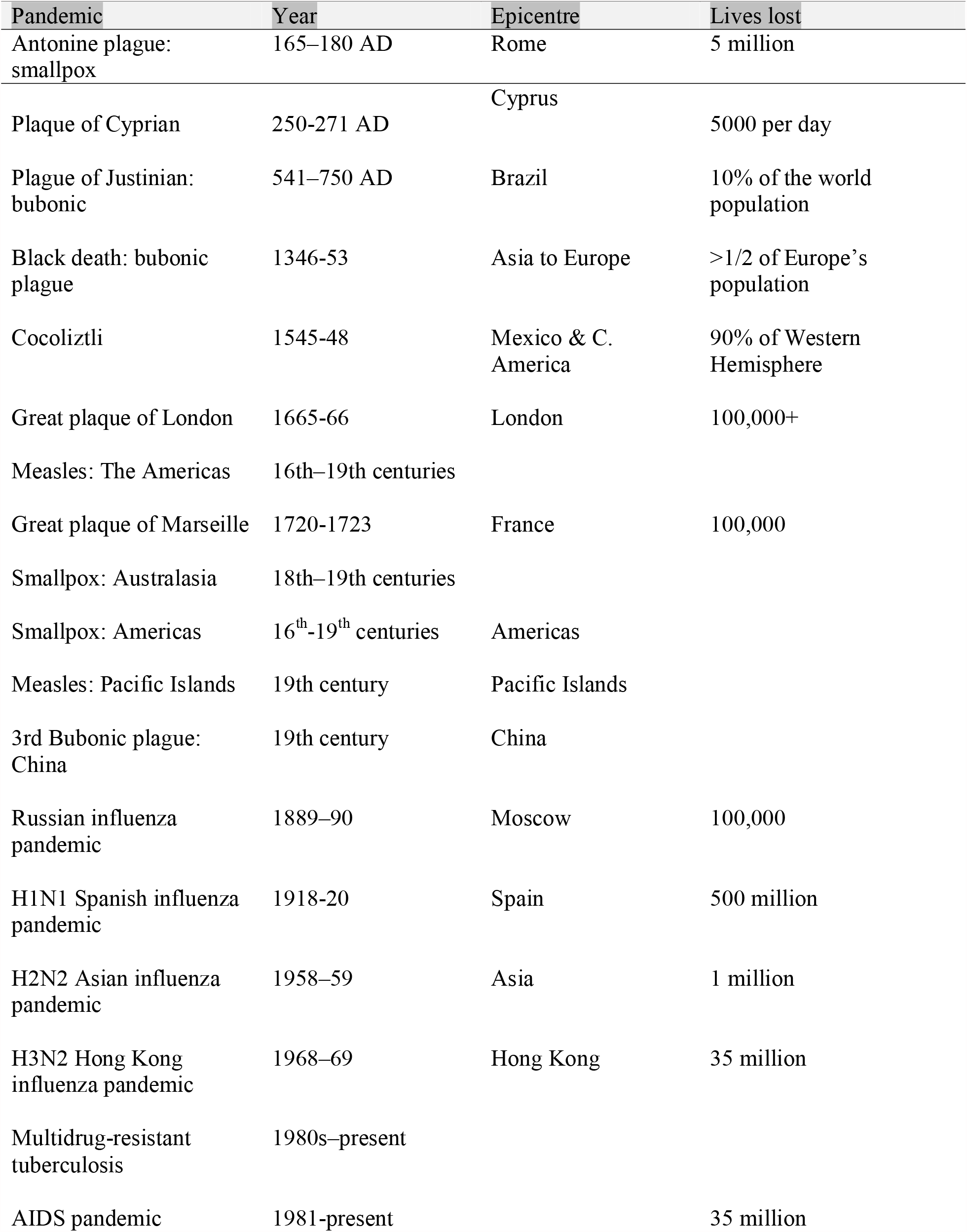
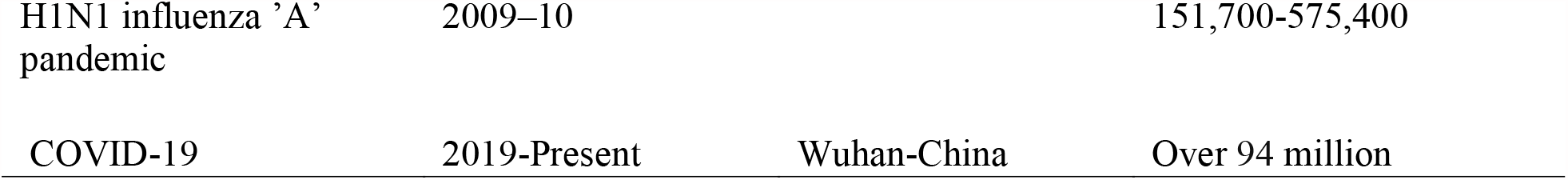
History of global pandemics, mortality, and epic centres

But there are some significant differences between the 2009 H1N1 swine flu and 2019 novel coronavirus (nCOVID-19) and pathophysiologically. By all serious estimates, COVID-19 is going to be a major killer.” Before the world experienced the 2009 H1N1 swine flu pandemic there was the first H1N1 Spanish flu in 1918 which remains the deadliest pandemic in the human history ^23^. The 2009 swine flu pandemic was caused by a new strain of H1N1 that originated in Mexico in 2009. From spring 2009 to June 2009 WHO declared it a pandemic. Comparatively, from April 2009 to April 2010, the wine flu was associated with 12,500 deaths (mortality rate of about 0.02%), and over 274,000 hospitalizations out of the 60.8 million cases ^24^. The mortality rate for COVID-19 is presently much higher, around 2.13%. This is because, most strains of flu viruses, including those that cause seasonal flu, cause the highest percentage of deaths in people ages 65 and older. But in the case of the H1N1, older people seemed to have already built up enough immunity and were not much affected compared to COVID-19 ^25^.

### 2.2 COVID-19 Pathophysiology in humans

A new novel coronavirus-induced pneumonia COVID-19 (SARS-CoV-2) first appeared in Wuhan, China in December 2019 ^25^. From its outbreak till date, it has spread to several countries globally ^26^. As of 20^th^ July, 2021, there were over 191 million confirmed cases of COVID-19 reported (in accordance with the applied case definitions and testing strategies in the affected countries), including 4.1 million deaths and this COVID-19 is steadily growing by human-to-human transmission ^26^.

The pathogenesis of Covid-19 is still not clear. Little is known about the pathogenesis of coronavirus disease in humans. Coronavirus disease (COVID-19) is caused by SARS-COV-2 which is a potentially fatal disease that is of great global public health concern. Patients with COVID-19 show similar symptoms of SARS-CoV and MERS-CoV which include fever, fatigue, dry cough, dyspnea, myalgia, normal or decreased leukocytes counts, proinflammatory cytokines and radiographic evidence of pneumonia ^27,28^. Severe pneumonia characterized by interstitial pneumonia, in which there is alveolar fibrosis, as a consequence of congestion, oedma and remodeling of lung parenchyma ^29^ and necrotizing alveolitis/bronchiolitis; characterized by foci necrosis of the epithelium, and secretions into the lumina is also seen in COVID-19 patient. The above mechanisms impair gaseous exchange mechanisms, leading to hypoxia/hypoxaemia and consequently, severe respiratory distress syndrome and multiorgan failure ^26,30^.

The virus is mainly spread during close contact and through respiratory droplets. The virus accesses host cells through the enzyme ACE2 (figure 2), which is most abundant in the type II alveolar cells of the lungs. ACE2, found in the lower respiratory tract of humans, is identified as cell receptor sites for SARS-CoV ^31^ and regulates both the cross-species and human-to-human transmission ^32^. The lungs are the therefore the most target organ of infection by the COVID-19 virus. The virus uses a special surface glycoprotein called a “spike” (peplomer) (figure 2) to connect to ACE2 and enter the host cell. Isolated from the bronchoalveolar lavage fluid (BALF) of a COVID-19 patient, Zhou *et al*., (^33^) have confirmed that the SARS-CoV-2 uses the same cellular entry receptor, ACE2, as SARS-CoV. The viral peplomer is pathogenic, inducing host immune response ^34^. The virion S-glycoprotein on the surface of coronavirus (figure 2) can attach to the receptor, ACE2 on the surface of human cells ^35^. ACE2 protein presents in abundance on lung alveolar epithelial cells and enterocytes of small intestine remarkably which may help understand the routes of infection and disease manifestations. Based on current epidemiological investigation, the incubation period is 1**–**14 days, mostly 3**–**7 days. And the COVID-19 is contagious during the latency period ^36^. It is highly transmissible in humans, especially in the elderly and people with underlying diseases.

**Figure 2:**
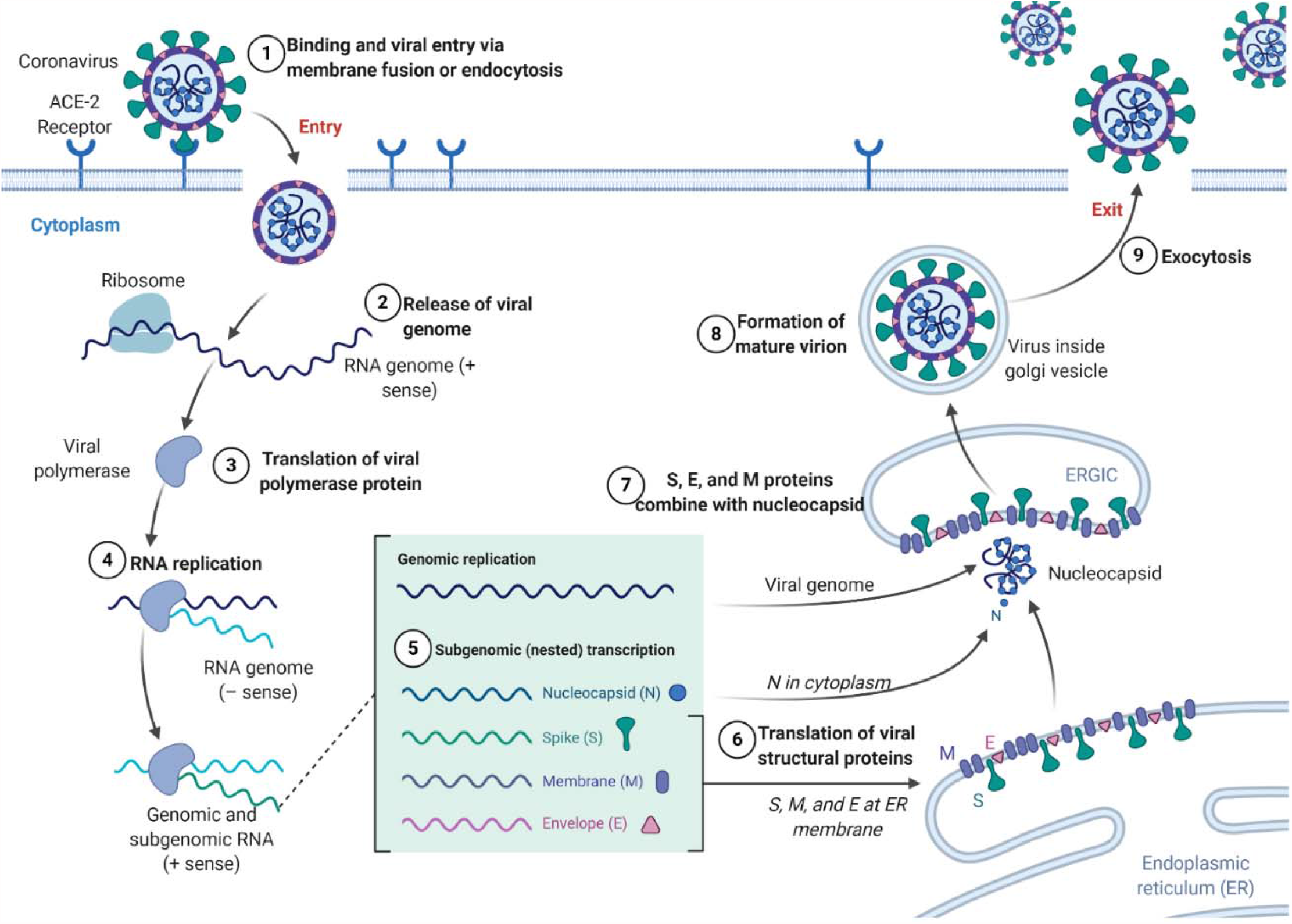
The life cycle of CoV in host cells. (Created with biorender.com)

The first event after internalization into host tissue is viral uncoating followed by translation of the viral genomic RNA to produce a virus-specific RNA-dependent RNA polymerase and various viral proteins shown in figure 2 ^37^. Clinically, patient presents with difficulty in breathing (a direct result of airway congestion and obliterating alveolitis), chest tightness, fever, sore throat (due to activation and proliferation of tonsilar lymph nodes). Other extrapulmonary manifestations are due to severe inflammatory response syndrome.

The virus spike protein (peplomer) acts as an exogenous pyrogen which upset the hypothalamic temperature regulatory set point (increases the set point) via cyclooxygenase dependent prostalglandins E2 production. Inflammatory cells such as lymphocytes and polymorphs have also been reported to be involved in the pneumonia caused by coronavirus.

The corona viral particle (COVID-19) attaches to the cellular receptor angiotensin-converting enzyme 2 (ACE2), releases its viral genomic RNA into the host cell and it is translated into proteins necessary for the assembly of new virions within the host cell. Key; S: spike, E: envelope, M: membrane, N: nucleocapsid. PP: polyproteins, ORF: Open reading frame, CoV: coronavirus. Adapted from ^38^.

### 2.3 Safety and efficacy of ginseng as an alternative medicine for disease control

Ginseng has been attributed with a large number of therapeutic effects including antioxidative, anti-inflammatory, vasorelaxant, antiallergic, antidiabetic, and anticancer effects ^39^. Ginseng extract alone or in combination with other herbs have been employed by herbal therapeutic and preventive medicine in the treatment of a number of medical illnesses ^11^. Ginseng is well known for its immune modulating effect and has been utilized in immune homeostasis and promoting resistance to illness or microbial infection through the immune system effects ^39^. Of particular importance is its use in respiratory infections such as chronic obstructive pulmonary disease, influenzas like illnesses and other respiratory tract infections ^12, 13, 14, 40^. The main active component of Ginseng that is responsible for its pharmaceutical activities and drug interactions are the Ginsenosides ^41^. There are many types of Ginsenoside with type Rg3 being one of the most important functions on lung diseases. It possesses anti-inflammatory, anti-tumour and anti-fatiguing properties. ^42^. Ginseng has been generally believed to be protective against pulmonary diseases ^43^. However, the studies on the efficacy of ginseng in the treatment of pulmonary diseases have been inconclusive due to contradicting evidences from different clinical trials with some studies establishing no clinical significance in alleviation of respiratory symptoms in clinical trials involving ginseng and placebos ^40,42^. Moreover, other studies have suggested promising pulmonary function and quality of life in patient with chronic obstructive pulmonary disease with the use of ginseng ^44,45^.

Ginseng has been shown to be relatively safe and well tolerated with no significant adverse reaction noted when compared with placebo during clinical trials ^39^. In addition, drug interaction with Ginseng appears to be rare as drug interaction studied have been inconclusive and have largely yielded negative results or results that suggest only a weak interaction ^46^.

### 2.4 Clinical characteristics

## 3. Methods

### 3.1. Data sources and selection

We conducted a comprehensive systematic literature search by employing the 6 electronic literature databases MEDLINE [15], PUBMED [17], SCOPUS [65], GOOGLE SCHOLAR [439], WEB OF SCIENCE [42] and the COCHRANE LIBRARY [33] from 1^st^ April, 2020 to 27^th^ April, 2020. Furthermore, Medical Subject Heading (MeSH) search was done at National Library of Medicine to create the MeSH terms. Secondary, additional manual search were done by following the relevant reference list of the selected papers. In addition, leading companies with trials on ginseng for respiratory diseases were contacted via email.

Studies in each language were screened using the following inclusion criteria: (1) human subjects, (2) use of a control procedure, (3) subjects randomized among treatment conditions, and (4) mono-preparation tests of Panax ginseng or P. quinquefolium.

### Search strategy

The database search was conducted in the month of April 2020 from the databases using the MESH terms linked by a Boolean search operators like ‘OR’ for the key MESH terms ‘Ginseng’’Panax Eleutherococcus’, ‘Eleuthero’, ‘Acanthopanax’, ‘Oplopanax’, ‘Echinopanax’ ‘AND’ ‘Covid-19’, ‘Coronavirus’, ‘Corona virus’, ‘Virus infection’, ‘Viral infection’, ‘Acute respiratory’, ‘Respiratory disease’, ‘Respiratory illness’, and other key words like ‘Trial’, ‘clinical trial’, ‘Randomi’, ‘Controlled study’, ‘Double blind’ and ‘Placebo’ and ‘AND ‘aged’.

An initial assessment using the inclusion criteria was made by reading abstracts. Articles that appeared to meet the criteria were then read in full by two authors, who then discussed the articles and made the decision to include or exclude them.

### 3.2. Data extraction and methodological quality assessment

Two authors extracted data from the articles using a standardized, predefined method that considered trial methods, study design, patient characteristics, type of ginseng, outcomes, and side effects. We used the Jadad scale to evaluate the quality of clinical trials ^59^. Points were awarded depending on the description of randomization, double-blinding, and appropriate/inappropriate methods, including withdrawals and dropouts. On a five-point scale, trials with three or more points were considered high quality. Discrepancies were settled through discussions involving two authors.

### 3.3. Review process

The RCTs were heterogeneous with respect to ginseng species or variety, indications, dose, participant characteristics, and outcome measures. The outcomes of some studies, however, were poorly presented. Therefore, we decided not to pool the data statistically, but performed a qualitative review instead. We reviewed RCTs to formulate conclusions on the effectiveness of ginseng for the following indications: glucose metabolism, physical performance, sexual function, psychomotor function, cardiac function, pulmonary disease, and cerebrovascular function. This method consisted of four levels of evidence on the methodological quality and outcome of the studies as follows: level 1, strong evidence, from generally consistent findings of multiple relevant, high-quality RCTs; level 2, moderate evidence, from generally consistent findings of one relevant, high-quality RCT and one or more relevant, low-quality RCTs; level 3, limited evidence, from generally consistent findings of multiple relevant, low-quality RCTs; and level 4, inconclusive evidence, from only one relevant, low-quality RCT, no relevant RCTs, or RCTs with conflicting results. “Generally consistent” was defined as two-thirds or more of the studies having the same result (positive or negative), and “multiple” was defined as more than one.

## 4. Results

Final data analyzed Our searches identified 596 potentially relevant studies, of which 5 trials met our inclusion criteria (Figure 3).

**Figure 3:**
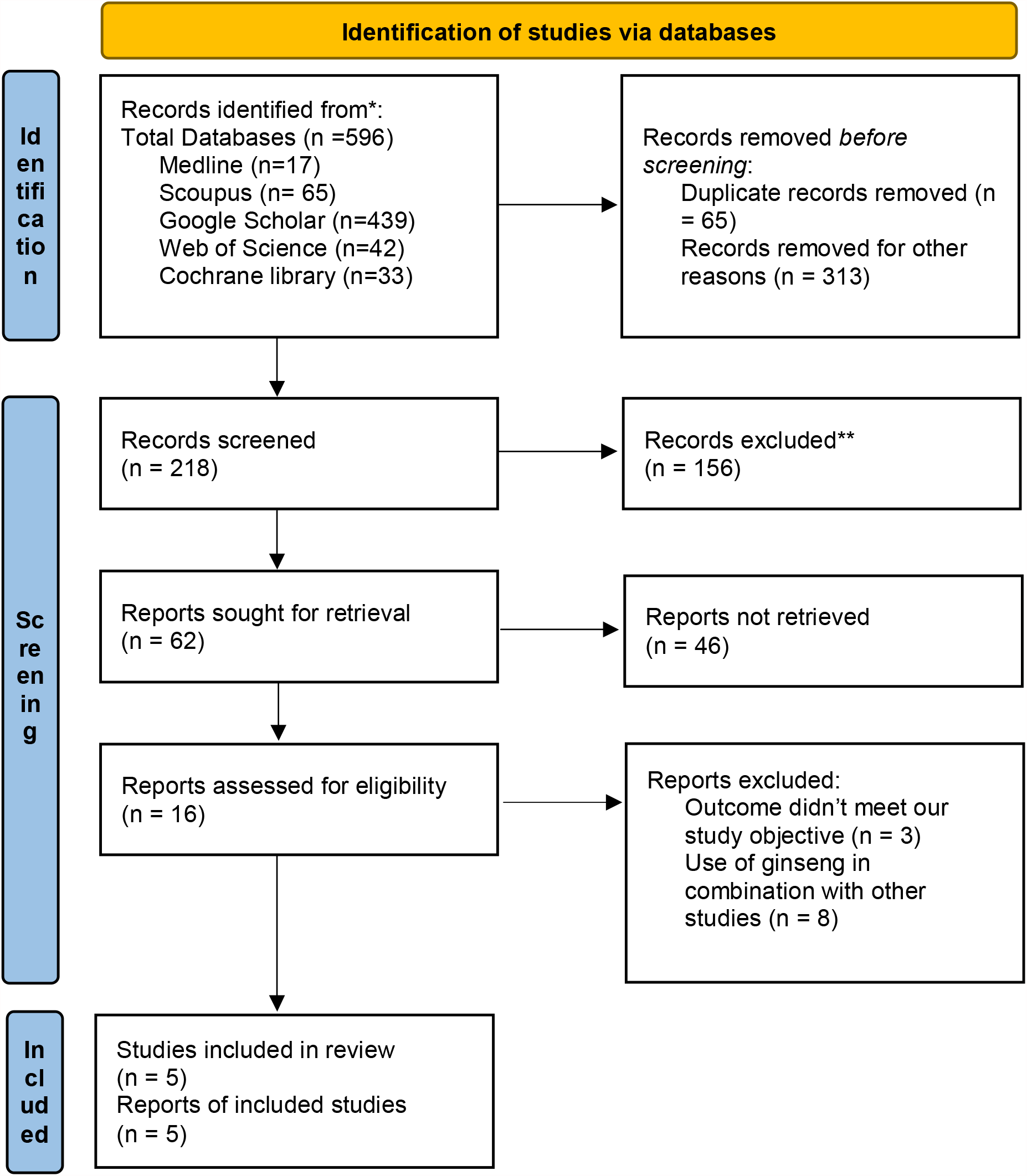
Updated 2020 PRISMA flow diagram for systematic review and meta-analysis; http://www.prisma-statement.org/.

The key data from all the included RCTs are summarized in Tables 4–6.

### 3.2. Description of studies and clinical questions

Of the 5 trials, 3 originated in Canada, 1 were in the United States, 1 were in South Korea. The clinical variables investigated were as follows: assessment of the effect of ginseng on acute respiratory diseases, prevention respiratory diseases.

**Table 2:**
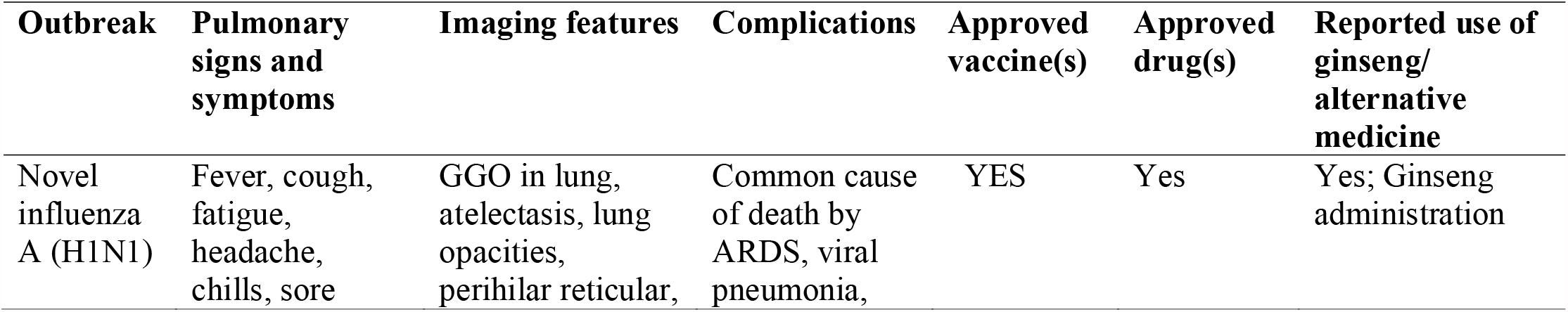
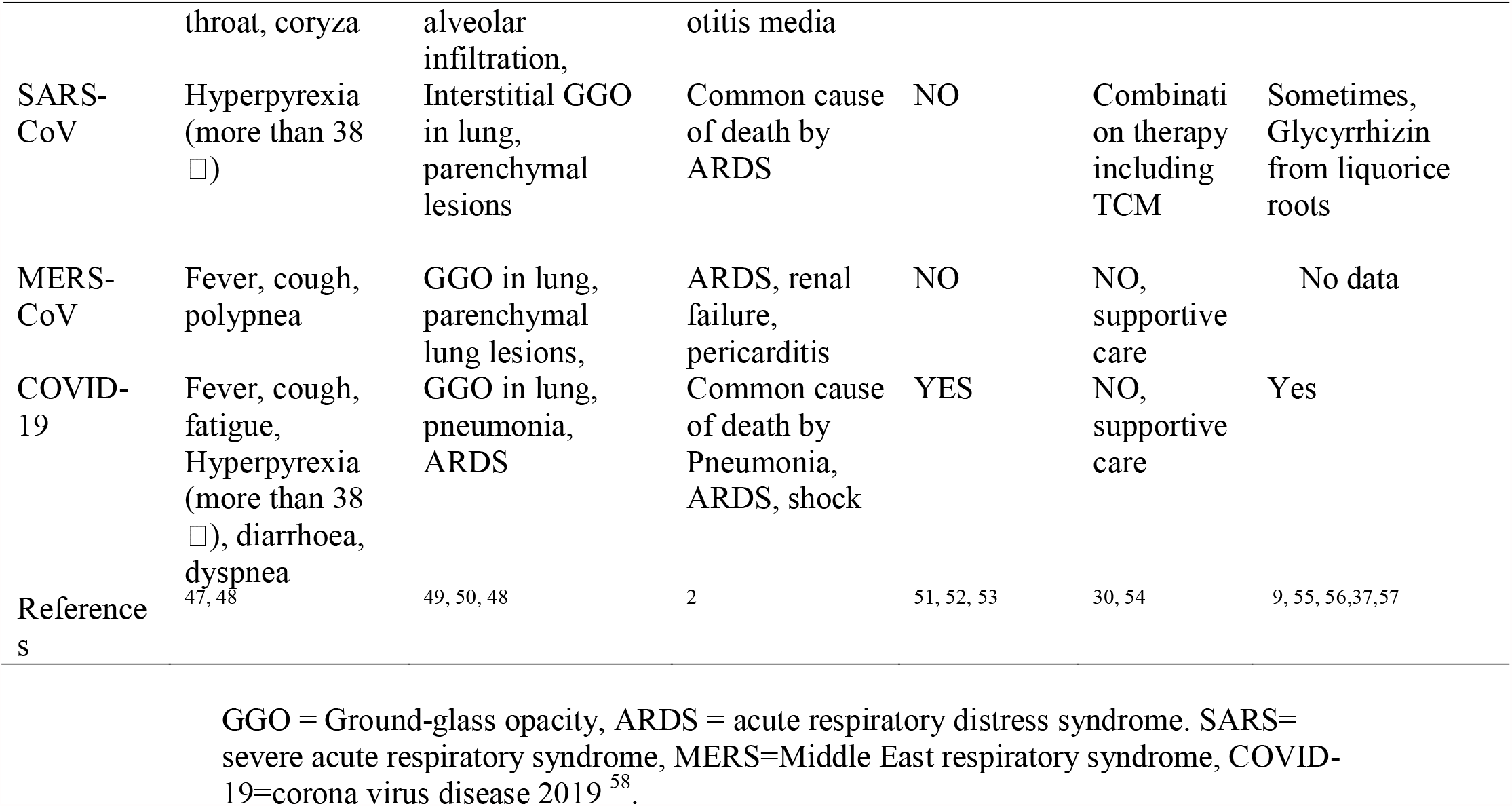
Clinical characteristics of SARS, MERS, H1N1 and COVID-19

**Table 4:**
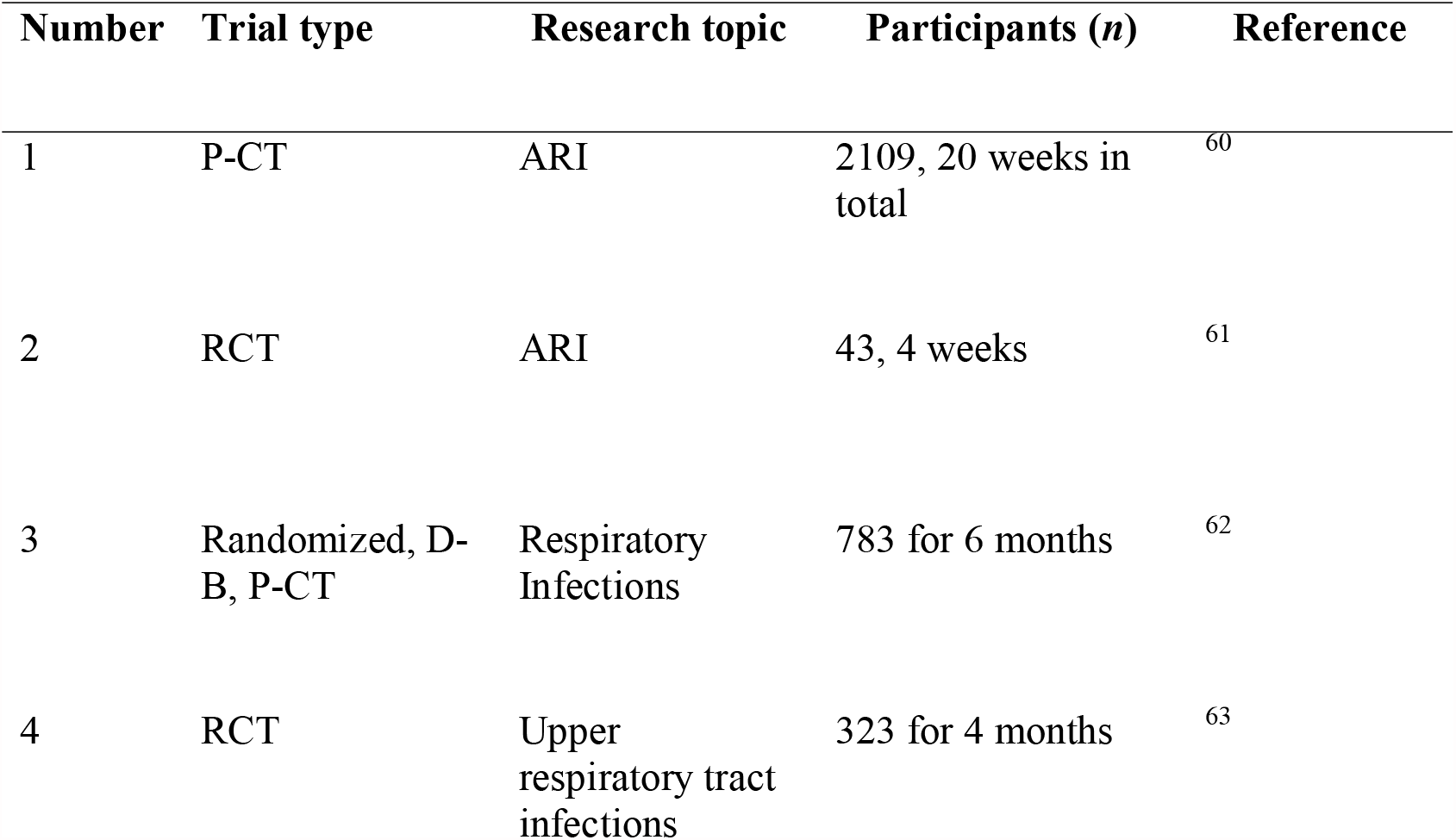
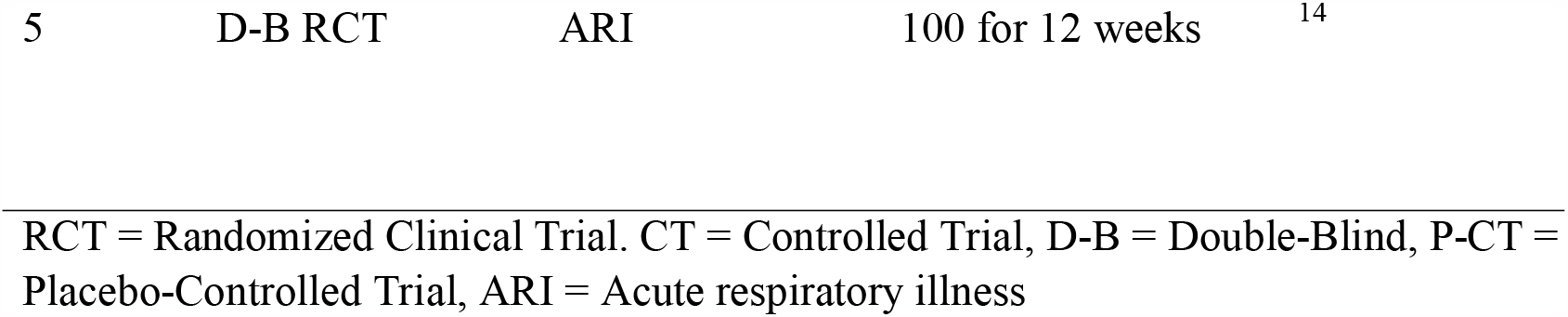
Summary of selected references.

## Meta-Analysis

The fixed effect and the random effect models gave similar values, and in these results, we present the random effect model results.

The meta-analysis study was carried out using the comprehensive meta-analysis package version 3. The pooled effect size for the Random Effect Model (REM) was 0.625 with lower-upper limit of (0.473-0.825) and *p* = 0.001) which means that ginseng significantly decreased the incidence of acute and upper respiratory infection by 38 %. This indicate that the participants who received the ginseng product, 38 % were protected from getting the infection and were at less risk compared to the placebo group. The p-value was less than 0.05 and indicates that the observed pooled effect was not due to chance. Again, the infections in the dosed group were less severe and it was observed that participant receiving the ginseng products recover about 3-days shorter (8.7 days) than the placebo (11.1 days) ^63, 56, 62^.

### Quality control check of the meta-analysis

Publication bias was evaluated with the funnel plot. The observation sows that there was less bias in the publication figure 5. The funnel plot was symmetrical, indicating free publication bias.

**Figure 4:**
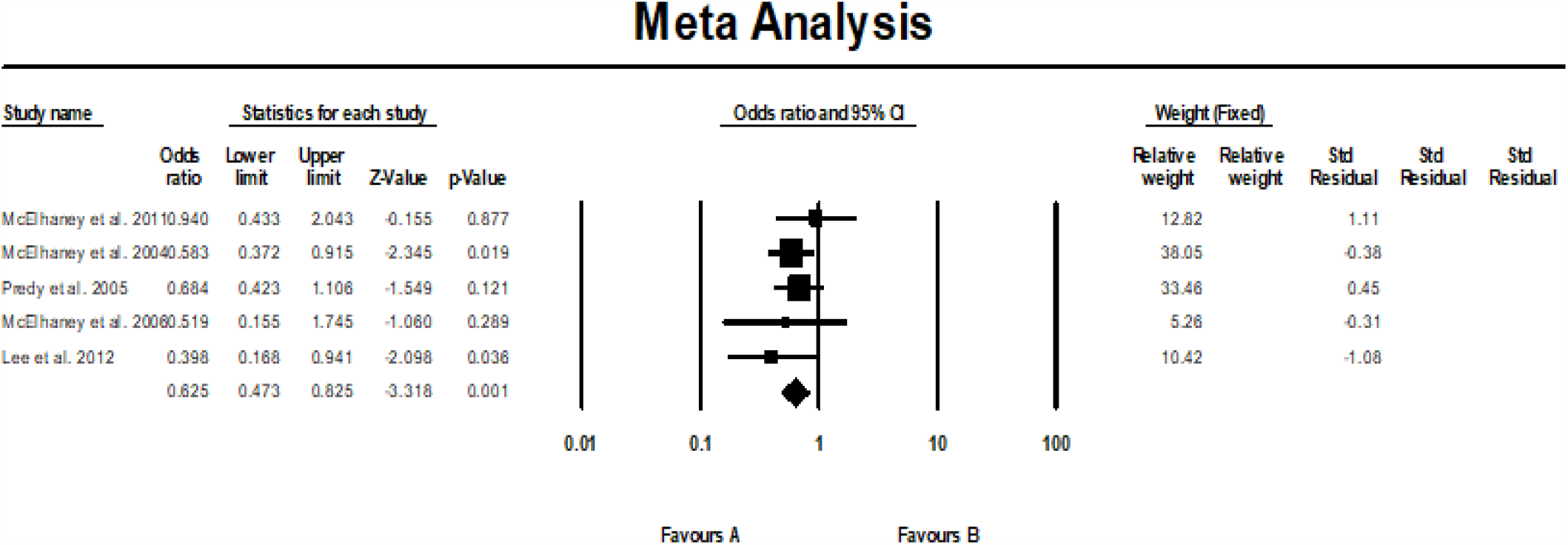
Meta-analysis results.

**Figure 5:**
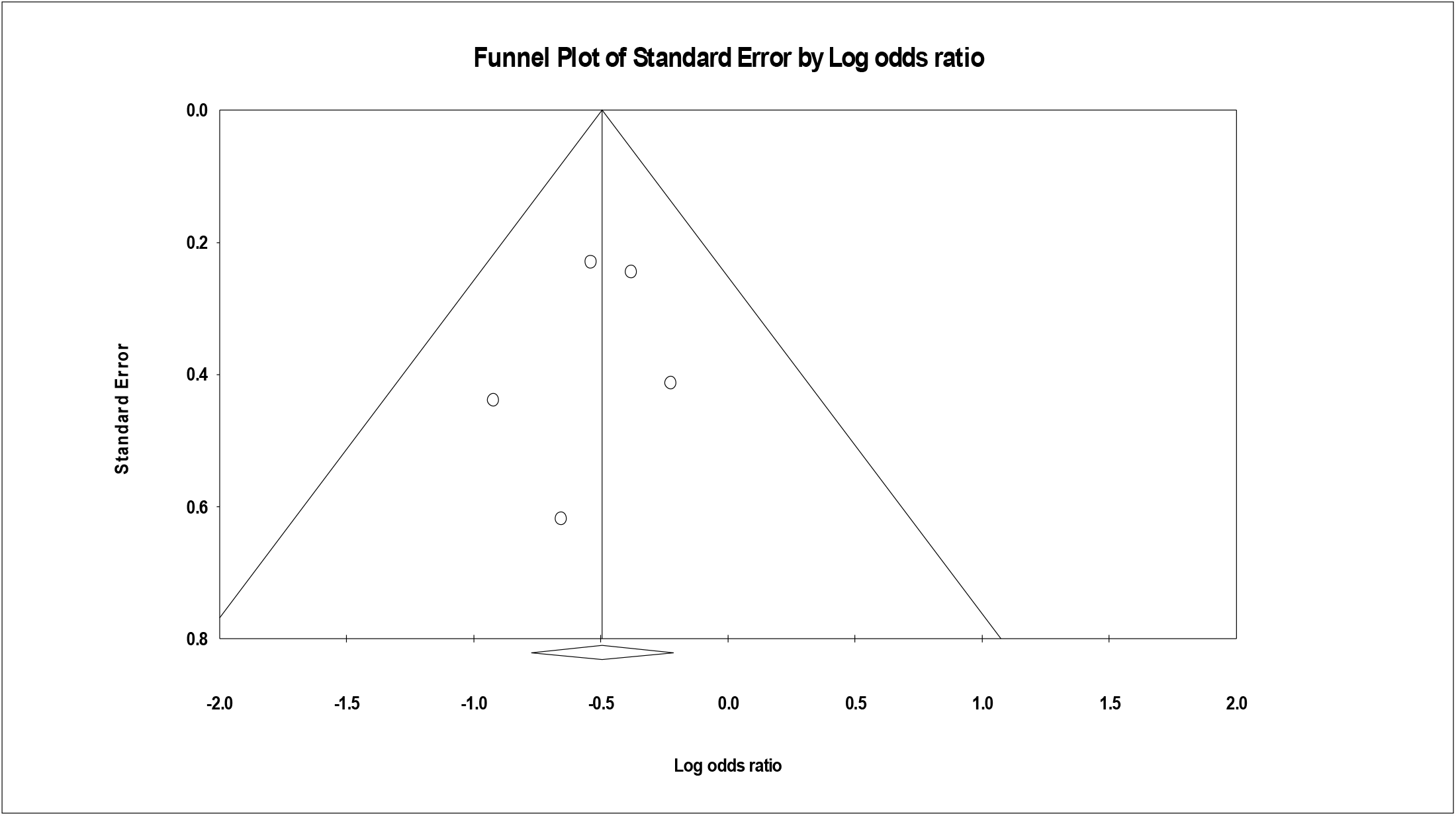
Funnel plot.

### The heterogeneity of the meta-analysis

Tau square (I^2^) of 0.00 % indicating no perceived between study variances as confirmed by the p-value for heterogeneity of greater than 0.05 (0.656); similar population sets. It can therefore be considered that the data was homogenous. In addition, the classical measure of heterogeneity which is Cochran’s Q-value for heterogeneity test which was 2.436 (df=4) indicating sufficiently homogeneous and a reliable result.

## 4.0 Discussion

Ginseng is a medicinal plant that has been used in medical practices for more than 2,000 years. In modern medical practice, ginseng has been used as an active substance in the treatment of disease and infections. For example, German Commission approved the use of ginseng as a tonic for reducing stress related to fatigue and declining sexual capacity ^64^. In addition, ginseng was approved by WHO in 1999 to enhance recovery.

Ginseng is believed to have a broad range of biological activities including anti-inflammatory, antioxidant and anti-tumor actions ^65^. Ginseng extracts has been shown to reduce to reduce the impacts of H1N1 infections ^42^. In a cell-based plaque assay study conducted by Kim et al. ^42^, oral administration of ginseng extract reduces the impact of H1N1 infection in mice. Plaque based assay can be used to determine the number of plaque forming units in a virus sample. This assay is effective in determining virus concentration in terms of infectious dose. Kim et al. ^42^ suggested that ginseng extract can be used in combination with conventional medicine in the treatment of H1N1 infections and coronaviruses.

Predy et al. ^63^ conducted an efficacy study using ginseng extracts in preventing upper respiratory tract infections in a randomized controlled trial. From their study, ingestion of a poly-furanosyl-pyranosyl-saccharide–rich extract of the roots of North American ginseng in a moderate dose over 4 months reduced the mean number of colds per person, the proportion of subjects who experienced 2 or more colds, the severity of symptoms and the number of days of cold symptoms among dosed group were less than the control group.

Ginseng extracts has been used in few clinical trials. Published clinical trial results showed that ginseng is safe at various dosages and can be effective in relieving the symptoms and reducing the risk and duration of colds and flu. Taken together with conventional medicine, these findings support the efficacy of ginseng as a therapeutic and prophylactic agent for respiratory infections.

In this review we summarized the systematic assessment of 5 double-blind RCTs on the effectiveness of Ginseng in the treatment of acute respiratory illness and upper respiratory tract infection. Importantly, only 5 of all identified publication met our inclusion criteria (Fig. 3). A total of 5 out of 596 identified publications were selected and only 3 out of the 5 selected studies were considered to have good methodological quality because these 3 studies have score value of greater that 3 points on Jadad scoring system (Table 3 - 5). A total of 3 out of the 5 selected studies investigated the effectiveness of ginseng on ARI treatment while 2 studies investigated the effectiveness of ginseng on URI treatment.

It is noteworthy that there have been various publications that have claimed that Ginseng is efficient in improving immune responses, effective in treating diseases such as cancer, cardiovascular disease and treating acute respiratory diseases. However, most of these claims are based on uncontrol and nonrandomized clinical studies ^66^. In order to streamline our identified publications to good quality studies, with good methodology and proper controls, we screened the identified articles using inclusion criteria (Fig. 3).

In combination with other herbs, Ginseng has previously been used to treat chronic respiratory diseases and upper respiratory tract infections ^62^. In addition, Ginseng has also been useful in the treatment of influenza like illness and respiratory tract infection ^14^. However, to the best of our knowledge its effectiveness in treating symptoms associated with COVID-19 has not be explored. In this systematic review we evaluated the research evidence currently available to access the effectiveness of ginseng in the treatment and prevention of ARI and URI.

Three out of the five reviewed RTCs showed that Ginseng reduces the risk and duration of acute respiratory illness with no accompanying significant adverse event. In RTC conducted by McElhaney et al ^60^, where they compared the effectiveness of American ginseng, CVT-E002, with placebo in institutionalized elderly people between 2000 and 2001. Their result showed that CVT-E002 has potential to prevent ARI ^60^. Also, in a similar RTC study conducted by McElhaney et al. ^61^, where they tested the effectiveness of ginseng in preventing ARI COLD-fX (CVT-E002) on elderly people, their result showed that COLD-fX reduce the risk and duration of ARI symptoms ^60^. Also, Lee et al ^56^, investigated the effectiveness of Korean Red Ginseng (KRG) on ARI treatment using 100 volunteers. They concluded that Korean Red Ginseng (KRG) has tendency to prevent subject from contracting ARI and can also reduce the duration of ARI symptoms 56.

In addition, in a separate RTCs studies by McElhaney et al and Predy et al ^62,63^, where they investigated the efficacy of ginseng in treatment of URI in healthy adults. They both concluded that ginseng reduced the severity and duration of URI symptoms ^62,63^.

While there is an ongoing race to develop an effective drug and/ vaccine to cure and prevent the spread of Covid-19. It is worth noting that, in the absence of an effective vaccine or antiviral drug for treatment of COVID-19 patients, health care professionals have adopted a supportive care strategy which involves effort to alleviate the symptoms of COVID-19 patients are mainly respiratory symptoms such as fever, dry cough, sore throat and sputum production.

As a result, the findings from these RCTs strengthened the claim that Ginseng is effective in treating ARI and URI. In addition, all the five papers reviewed did not report a significant adverse event that is related to Ginseng usage during the period of these studies (Table 6).

**Table 5:** Methodological quality of included studies according to Jadad et al. (Jadad et al., 1996) and Schultz et al. (Schulz et al., 1995).

| Reference | Des. as randomized | Randomization well des. and appropriate | Outcome assessment blinded | Blinding well Des. and appropriate | Description of Withdrawals/drop-outs | Total Jadad score | Allocation concealment |
| --- | --- | --- | --- | --- | --- | --- | --- |
| McElhaney JE *et al.* (2004)<sup>60</sup> | Yes | No | Yes | No | Yes | 3/5 | Unclear |
| McElhaney JE *et al.* (2006)<sup>61</sup> | Yes | Yes | Yes | Yes | No | 4/5 | Clear |
| McElhaney *et al.*, (2011)<sup>62</sup> | Yes | Yes | Yes | Yes | Yes | 5/5 | Clear |
| Predy *et al.*, (2005)<sup>63</sup><br>Canada | Yes | Yes | Yes | Yes | Yes | 5/5 | Clear |
| Lee *et al.*, 2012<sup>56</sup> | Yes | No | Yes | No | Yes | 3/5 | Unclear |

Key: Des = Described,

**Table 6:**
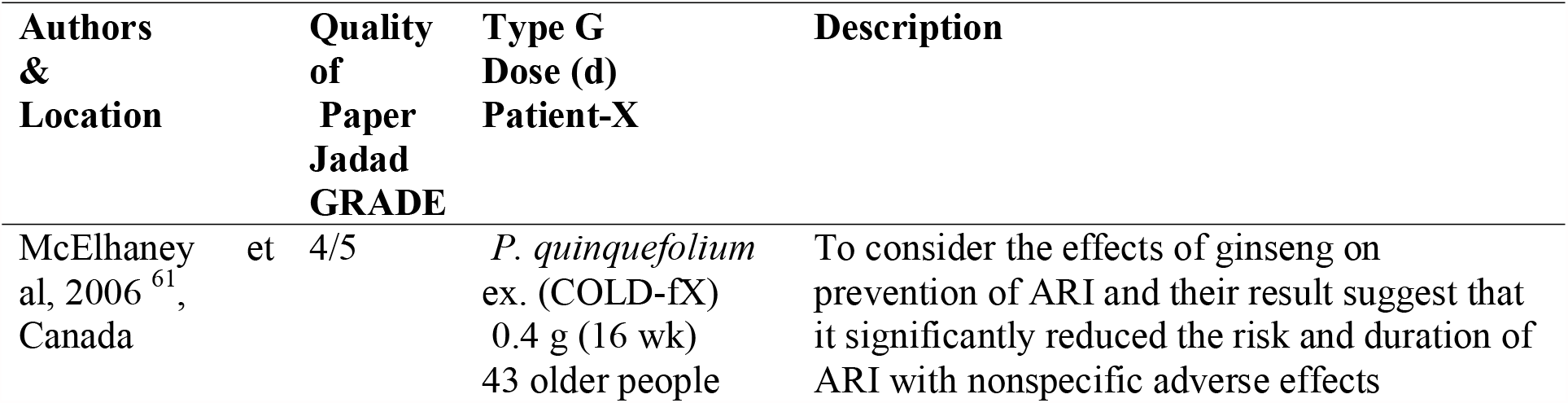
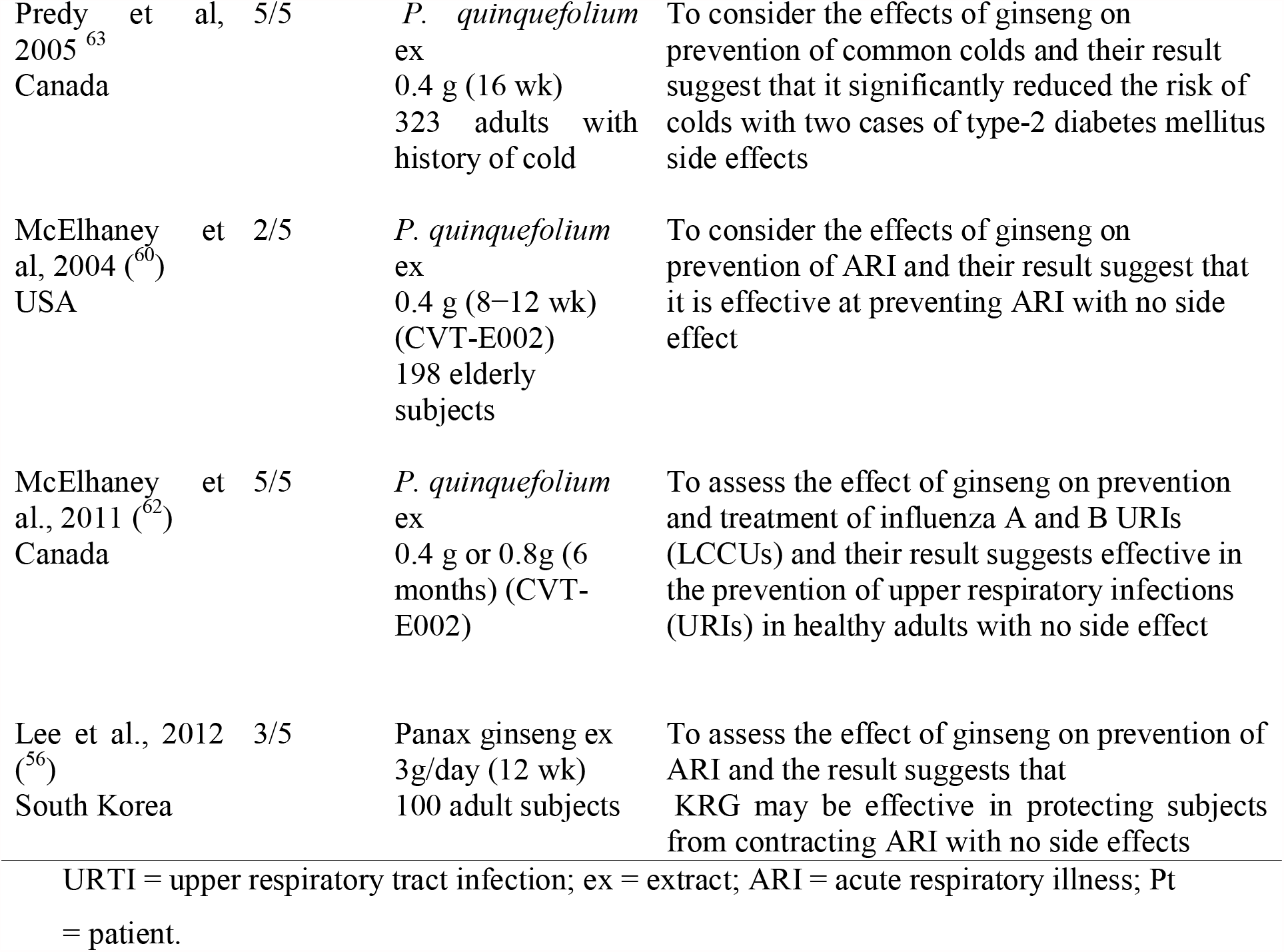
Focus of the selected studies.

While previous studies have emphasized the beneficial effect of Ginseng for various therapeutic purposes. The five RCTs reviewed in this paper have provided a platform for consideration of Ginseng for use in supportive care of COVID-19 patients. We hope this review serves as a wake-up call for consideration of Ginseng in treatment of COVID-19.

The result from the meta-analysis confirms the applicability of ginseng to significantly prevent acute respiratory viral infections including coronavirus disease in humans with a reduction rate of over 60%. The finding is reliable as the studies reviewed had low degree of bias and very homogeneous with I2 of 0.00 and Cochrane’s q-value of 2.4.

## 5.0 Conclusion

The swift emergence of new infectious virus and drug-resistant variants has limited the availability of effective antiviral agents and vaccines. Thus, the development of broad-spectrum antivirals and immunomodulating agents that stimulate host immunity and improve host resilience is essential. Acute Respiratory illnesses (especially among adult population) require significant medical intervention in the primary health-care setting and it should be taken as a matter of urgency, irrespective of gender and age. In spite of the worldwide expansion of the use of ginseng at various dosages over the years, some clinical trials have shown that ginseng has the potential to prevent acute and upper respiratory illnesses, by relieving the symptoms and reducing the risk and duration of the manifestation of different respiratory viruses causing illness (SARS etc.).

As the race to develop an efficient drug / vaccine to prevent and treat the spread of COVID-19 continues, this study has been able to reveal the effectiveness of the use of ginseng over the years in treating and alleviating the symptoms of upper and acute respiratory illnesses and further studies into the applicability of ginseng in coronavirus disease prevent and severity reduction is warranted. Ginseng was also observed that the dosed but infected group had less severe infection, shorter sickness duration with faster recovery times than the placebo ^63, 56, 62^. Although it is generally believed that ginseng is protective against pulmonary diseases, more research work and clinical trials which can further reveal its benefits for therapeutic purposes, including the treatment of both young and old COVID-19 patients are needed. Also, it will be prudent to further examine factors that may precipitate adverse events as a result of frequent or prolonged ingestion of regulated ginseng extracts.

## Data Availability

All relevant data has been included with the submission

## Appendix A: Search strategies with the Boolean operators and MeSH Terms as used in Web of Science

(TITLE-ABS-KEY(ginseng OR “jen shen” OR ninjin OR renshen OR “ren shen” OR schinseng OR shinseng OR panax OR eleutherococcus OR eleuthero OR acanthopanax OR ciwujia OR oplopanax OR echinopanax)) AND (TITLE-ABS-KEY(“covid 19” OR coronavirus* OR “corona virus*” OR “virus infection*” OR “viral infection*” OR “acute respiratory” OR “respiratory disease*” OR “respiratory illness*” OR “respiratory infection*” OR “respiratory syndrome*” OR “respiratory tract disease*” OR “respiratory tract illness*” OR “respiratory tract infection*” OR “respiratory tract syndrome*” OR sars OR mers OR “pulmonary disease*” OR “pulmonary illness*” OR “pulmonary infection*” OR “pulmonary syndrome*” OR “lung disease*” OR “lung illness*” OR “lung infection*” OR “lung syndrome*”)) AND (TITLE-ABS-KEY(trial* OR randomi* OR “controlled stud*” OR “double blind” OR placebo*)) AND (TITLE-ABS-KEY(aged OR aging OR elder* OR older OR senior* OR “old person*” OR “old people” OR “old population*”))

## References

1. WHO. Coronavirus. World Health Organisation. Published 2020. Accessed April 27, 2020. https://www.who.int/westernpacific/health-topics/coronavirus

2. WHO. WHO Director-General’s opening remarks at the media briefing on COVID-19 - 11 March 2020. Published 2020. Accessed April 27, 2020. https://www.who.int/dg/speeches/detail/who-director-general-s-opening-remarks-at-the-media-briefing-on-covid-19---11-march-2020

3. Dhama K, Sharun K, Tiwari R, et al. Coronavirus Disease 2019 &ndash; COVID-19. Published online March 1, 2020. doi:10.20944/preprints202003.0001.v1

4. Lu X, Zhang L, Du H, et al. SARS-CoV-2 Infection in Children. New England Journal of Medicine. 2020;382(17):1663–1665. doi:10.1056/NEJMc2005073

5. Amanat F, Krammer F. SARS-CoV-2 Vaccines: Status Report. Immunity. 2020;52(4):583–589. doi:10.1016/j.immuni.2020.03.007

6. Liu J, Manheimer E, Shi Y, Gluud C. Chinese Herbal Medicine for Severe Acute Respiratory Syndrome: A Systematic Review and Meta-Analysis. THE JOURNAL OF ALTERNATIVE AND COMPLEMENTARY MEDICINE. 2004;10(6):1041–1051.

7. Yang Y, Islam MS, Wang J, Li Y, Chen X. Traditional Chinese Medicine in the Treatment of Patients Infected with 2019-New Coronavirus (SARS-CoV-2): A Review and Perspective. Int J Biol Sci. 2020;16(10):1708–1717. doi:10.7150/ijbs.45538

8. Chen W, Lim CED, Kang H-J, Liu J. Chinese Herbal Medicines for the Treatment of Type A H1N1 Influenza: A Systematic Review of Randomized Controlled Trials. PLoS One. 2011;6(12). doi:10.1371/journal.pone.0028093

9. Luo H, Tang Q, Shang Y, et al. Can Chinese Medicine Be Used for Prevention of Corona Virus Disease 2019 (COVID-19)? A Review of Historical Classics, Research Evidence and Current Prevention Programs. Chin J Integr Med. 2020;26(4):243–250. doi:10.1007/s11655-020-3192-6

10. Scaglione F, Ferrara F, Dugnani S, Falchi M, Santoro G, Fraschini F. Immunomodulatory effects of two extracts of Panax ginseng C.A. Meyer. Drugs Exp Clin Res. 1990;16(10):537–542.

11. An X, Zhang AL, Yang AW, et al. Oral ginseng formulae for stable chronic obstructive pulmonary disease: a systematic review. Respir Med. 2011;105(2):165–176. doi:10.1016/j.rmed.2010.11.007

12. Barth A, Hovhannisyan A, Jamalyan K, Narimanyan M. Antitussive effect of a fixed combination of Justicia adhatoda, Echinacea purpurea and Eleutherococcus senticosus extracts in patients with acute upper respiratory tract infection: A comparative, randomized, double-blind, placebo-controlled study. Phytomedicine. 2015;22(13):1195–1200. doi:10.1016/j.phymed.2015.10.001

13. Vohra S, Johnston BC, Laycock KL, et al. Safety and tolerability of North American ginseng extract in the treatment of pediatric upper respiratory tract infection: a phase II randomized, controlled trial of 2 dosing schedules. Pediatrics. 2008;122(2):e402–410. doi:10.1542/peds.2007-2186

14. Ha K-C, Kim M-G, Oh M-R, et al. A placebo-controlled trial of Korean red ginseng extract for preventing Influenza-like illness in healthy adults. BMC Complementary and Alternative Medicine. 2012;12. doi:10.1186/1472-6882-12-10

15. Engel JP. Viral upper respiratory infections. Semin Respir Infect. 1995;10(1):3–13.

16. El Zowalaty ME, Järhult JD. From SARS to COVID-19: A previously unknown SARS-related coronavirus (SARS-CoV-2) of pandemic potential infecting humans – Call for a One Health approach. One Health. 2020;9:100124. doi:10.1016/j.onehlt.2020.100124

17. Chin AWH, Chu JTS, Perera MRA, et al. Stability of SARS-CoV-2 in different environmental conditions. medRxiv. Published online March 27, 2020:2020.03.15.20036673. doi:10.1101/2020.03.15.20036673

18. Arbo MD, Schmitt GC, Limberger MF, et al. Subchronic toxicity of Citrus aurantium L. (Rutaceae) extract and p-synephrine in mice. Regulatory Toxicology and Pharmacology. 2009;54(2):114–117. doi:10.1016/j.yrtph.2009.03.001

19. Nijsingh N, van Bergen A, Wild V. Applying a Precautionary Approach to Mobile Contact Tracing for COVID-19: The Value of Reversibility. Bioethical Inquiry. 2020;17(4):823–827. doi:10.1007/s11673-020-10004-z

20. Taubenberger JK. The Origin and Virulence of the 1918 “Spanish” Influenza Virus. Proc Am Philos Soc. 2006;150(1):86–112.

21. Kakodkar P, Kaka N, Baig MN. A Comprehensive Literature Review on the Clinical Presentation, and Management of the Pandemic Coronavirus Disease 2019 (COVID-19). Cureus. 2020;12(4):e7560. doi:10.7759/cureus.7560

22. Gössling S, Scott D, Hall CM. Pandemics, tourism and global change: a rapid assessment of COVID-19. Journal of Sustainable Tourism. 2021;29(1):1–20. doi:10.1080/09669582.2020.1758708

23. Erkoreka A. Origins of the Spanish Influenza pandemic (1918–1920) and its relation to the First World War. J Mol Genet Med. 2009;3(2):190–194.

24. Im K, Kim J, Min H. Ginseng, the natural effectual antiviral: Protective effects of Korean Red Ginseng against viral infection. Journal of Ginseng Research. 2016;40(4):309–314. doi:10.1016/j.jgr.2015.09.002

25. Zhou Y, Hou Y, Shen J, Huang Y, Martin W, Cheng F. Network-based drug repurposing for novel coronavirus 2019-nCoV/SARS-CoV-2. Cell Discov. 2020;6. doi:10.1038/s41421-020-0153-3

26. Wu D, Wu T, Liu Q, Yang Z. The SARS-CoV-2 outbreak: What we know. Int J Infect Dis. 2020;94:44–48. doi:10.1016/j.ijid.2020.03.004

27. Huang Y, Yang C, Xu X-F, Xu W, Liu S-W. Structural and functional properties of SARS-CoV-2 spike protein: potential antivirus drug development for COVID-19. Acta Pharmacol Sin. 2020;41(9):1141–1149. doi:10.1038/s41401-020-0485-4

28. Sharma A, Tiwari S, Deb MK, Marty JL. Severe acute respiratory syndrome coronavirus-2 (SARS-CoV-2): a global pandemic and treatment strategies. Int J Antimicrob Agents. 2020;56(2):106054. doi:10.1016/j.ijantimicag.2020.106054

29. Hosseini A, Hashemi V, Shomali N, et al. Innate and adaptive immune responses against coronavirus. Biomedicine & Pharmacotherapy. 2020;132:110859. doi:10.1016/j.biopha.2020.110859

30. Hilgenfeld R, Peiris M. From SARS to MERS: 10 years of research on highly pathogenic human coronaviruses. Antiviral Research. 2013;100(1):286–295. doi:10.1016/j.antiviral.2013.08.015

31. Jia HP, Look DC, Shi L, et al. ACE2 receptor expression and severe acute respiratory syndrome coronavirus infection depend on differentiation of human airway epithelia. J Virol. 2005;79(23):14614–14621. doi:10.1128/JVI.79.23.14614-14621.2005

32. Wan Y, Shang J, Graham R, Baric RS, Li F. Receptor Recognition by the Novel Coronavirus from Wuhan: an Analysis Based on Decade-Long Structural Studies of SARS Coronavirus. J Virol. 2020;94(7):e00127–20. doi:10.1128/JVI.00127-20

33. Zhou F, Yu T, Du R, et al. Clinical course and risk factors for mortality of adult inpatients with COVID-19 in Wuhan, China: a retrospective cohort study. Lancet. 2020;395(10229):1054–1062. doi:10.1016/S0140-6736(20)30566-3

34. Raj RS, Bonney EA, Phillippe M. Influenza, Immune System, and Pregnancy. Reprod Sci. 2014;21(12):1434–1451. doi:10.1177/1933719114537720

35. Walls AC, Park Y-J, Tortorici MA, Wall A, McGuire AT, Veesler D. Structure, Function, and Antigenicity of the SARS-CoV-2 Spike Glycoprotein. Cell. 2020;181(2):281–292.e6. doi:10.1016/j.cell.2020.02.058

36. Jin Y, Yang H, Ji W, et al. Virology, Epidemiology, Pathogenesis, and Control of COVID-19. Viruses. 2020;12(4):E372. doi:10.3390/v12040372

37. Poutanen SM. Human Coronaviruses. Principles and Practice of Pediatric Infectious Diseases. Published online 2018:1148–1152.e3. doi:10.1016/B978-0-323-40181-4.00222-X

38. Prajapat M, Sarma P, Shekhar N, et al. Drug targets for corona virus: A systematic review. Indian J Pharmacol. 2020;52(1):56–65. doi:10.4103/ijp.IJP_115_20

39. Lim SW, Luo K, Quan Y, et al. The safety, immunological benefits, and efficacy of ginseng in organ transplantation. Journal of Ginseng Research. 2020;44(3):399–404. doi:10.1016/j.jgr.2020.02.001

40. Shergis JL, Thien F, Worsnop CJ, et al. 12-month randomised controlled trial of ginseng extract for moderate COPD. Thorax. 2019;74(6):539–545. doi:10.1136/thoraxjnl-2018-212665

41. Li W, Yan M-H, Liu Y, et al. Ginsenoside Rg5 Ameliorates Cisplatin-Induced Nephrotoxicity in Mice through Inhibition of Inflammation, Oxidative Stress, and Apoptosis. Nutrients. 2016;8(9):566. doi:10.3390/nu8090566

42. Kim J-Y, Kim H-J, Kim H-J. Effect of Oral Administration of Korean Red Ginseng on Influenza A (H1N1) Virus Infection. Journal of Ginseng Research. 2011;35(1):104–110. doi:10.5142/jgr.2011.35.1.104

43. Xue CC, Shergis JL, Zhang AL, et al. Panax ginseng C.A Meyer root extract for moderate chronic obstructive pulmonary disease (COPD): study protocol for a randomised controlled trial. Trials. 2011;12:164. doi:10.1186/1745-6215-12-164

44. Shergis JL, D. YM, Zhang AL, et al. Therapeutic potential of Panax ginseng and ginsenosides in the treatment of chronic obstructive pulmonary disease. Complement Ther Med. 2014;22(5):944–953. doi:10.1016/j.ctim.2014.08.006

45. Iqbal H, Rhee D. Ginseng alleviates microbial infections of the respiratory tract: a review. J Ginseng Res. 2020;44(2):194–204. doi:10.1016/j.jgr.2019.12.001

46. Ramanathan MR, Penzak SR. Pharmacokinetic Drug Interactions with Panax ginseng. Eur J Drug Metab Pharmacokinet. 2017;42(4):545–557. doi:10.1007/s13318-016-0387-5

47. Poeppl W, Hell M, Herkner H, et al. Clinical aspects of 2009 pandemic influenza A (H1N1) virus infection in Austria. Infection. 2011;39(4):341–352. doi:10.1007/s15010-011-0121-9

48. Rothan HA, Byrareddy SN. The epidemiology and pathogenesis of coronavirus disease (COVID-19) outbreak. Journal of Autoimmunity. 2020;109:102433. doi:10.1016/j.jaut.2020.102433

49. Mollura DJ, Asnis DS, Crupi RS, et al. Imaging Findings in a Fatal Case of Pandemic Swine-Origin Influenza A (H1N1). American Journal of Roentgenology. 2009;193(6):1500–1503. doi:10.2214/AJR.09.3365

50. Si B, N MR, S AS, A K. Chest imaging features of patients afflicted with Influenza A (H1N1) in a Malaysian tertiary referral centre. Biomedical Imaging and Intervention Journal. 2010;6(4):e35.

51. Dubé E, Gilca V, Sauvageau C, et al. Canadian family physicians’ and paediatricians’ knowledge, attitudes and practices regarding A(H1N1) pandemic vaccine. BMC Res Notes. 2010;3(1):102. doi:10.1186/1756-0500-3-102

52. Saif LJ. Animal coronavirus vaccines: lessons for SARS. Developments in Biologicals. 2004;(119):129–140.

53. Al-Tawfiq JA, Memish ZA. An update on Middle East respiratory syndrome: 2 years later. Expert Review of Respiratory Medicine. 2015;9(3):327–335. doi:10.1586/17476348.2015.1027689

54. CDC. Antiviral Agents for the Treatment and Chemoprophylaxis of Influenza. Published 2011. Accessed April 30, 2020. https://www.cdc.gov/mmwR/preview/mmwrhtml/rr6001a1.htm

55. Tello C. Can Ginseng Help Fight COVID-19? SelfHacked. Published 2020. Accessed May 2, 2020. https://selfhacked.com/blog/ginseng-covid-19/

56. Lee C-S, Lee J-H, Oh M, et al. Preventive Effect of Korean Red Ginseng for Acute Respiratory Illness: A Randomized and Double-Blind Clinical Trial. J Korean Med Sci. 2012;27(12):1472–1478. doi:10.3346/jkms.2012.27.12.1472

57. Cinatl J, Morgenstern B, Bauer G, Chandra P, Rabenau H, Doerr HW. Glycyrrhizin, an active component of liquorice roots, and replication of SARS-associated coronavirus. Lancet. 2003;361(9374):2045–2046. doi:10.1016/s0140-6736(03)13615-x

58. Huan-Tian Cui Y-TL, Huan-Tian Cui Y-TL. Traditional Chinese medicine for treatment of coronavirus disease 2019: a review. Traditional Medicine Research. 2020;5(2):65–73. doi:10.12032/TMR20200222165

59. Olivo SA, Macedo LG, Gadotti IC, Fuentes J, Stanton T, Magee DJ. Scales to Assess the Quality of Randomized Controlled Trials: A Systematic Review. Physical Therapy. 2008;88(2):156–175. doi:10.2522/ptj.20070147

60. Mcelhaney JE, Gravenstein S, Cole SK, et al. A Placebo-Controlled Trial of a Proprietary Extract of North American Ginseng (CVT-E002) to Prevent Acute Respiratory Illness in Institutionalized Older Adults. Journal of the American Geriatrics Society. 2004;52(1):13–19. doi:10.1111/j.1532-5415.2004.52004.x

61. McElhaney JE, Goel V, Toane B, Hooten J, Shan JJ. Efficacy of COLD-fX in the prevention of respiratory symptoms in community-dwelling adults: A randomized, double-blinded, placebo controlled trial. Journal of Alternative and Complementary Medicine. 2006;12(2):153–157. doi:10.1089/acm.2006.12.153

62. McElhaney JE, Simor AE, McNeil S, Predy GN. Efficacy and Safety of CVT-E002, a Proprietary Extract of Panax quinquefolius in the Prevention of Respiratory Infections in Influenza-Vaccinated Community-Dwelling Adults: A Multicenter, Randomized, Double-Blind, and Placebo-Controlled Trial. Influenza Res Treat. 2011;2011:759051. doi:10.1155/2011/759051

63. Predy GN, Goel V, Lovlin R, Donner A, Stitt L, Basu TK. Efficacy of an extract of North American ginseng containing poly-furanosyl-pyranosyl-saccharides for preventing upper respiratory tract infections: a randomized controlled trial. CMAJ. 2005;173(9):1043–1048. doi:10.1503/cmaj.1041470

64. Park HJ, Kim DH, Park SJ, Kim JM, Ryu JH. Ginseng in Traditional Herbal Prescriptions. J Ginseng Res. 2012;36(3):225–241. doi:10.5142/jgr.2012.36.3.225

65. Block KI, Mead MN. Immune System Effects of Echinacea, Ginseng, and Astragalus: A Review. Integr Cancer Ther. 2003;2(3):247–267. doi:10.1177/1534735403256419

66. Vogler BK, Pittler MH, Ernst E. The efficacy of ginseng. A systematic review of randomised clinical trials. Eur J Clin Pharmacol. 1999;55(8):567–575. doi:10.1007/s002280050674

